# COVID-19 in Tunisia (North Africa): IgG and IgG subclass antibody responses to SARS-CoV-2 according to disease severity

**DOI:** 10.1101/2022.03.01.22271696

**Authors:** Chaouki Benabdessalem, Soumaya Marzouki, Wafa Ben Hamouda, Khaled Trabelsi, Mohamed Boumaiza, Sonia Ben Hamouda, Rym Ouni, Soumaya Bchiri, Amani Chaaban, Meriem Gdoura, Yousr Gorgi, Imen Sfar, Sadok Yalaoui, Jalila Ben Khelil, Agnes Hamzaoui, Meya Abdallah, Yosra Cherif, Stéphane Petres, Chris Ka Pun Mok, Nicolas Escriou, Sébastien Quesney, Koussay Dellagi, Jihene Bettaieb, Samia Rourou, Mohamed Ridha Barbouche, Melika Ben Ahmed

## Abstract

Coronavirus disease 2019 (COVID-19) expresses a wide spectrum of disease severity. We investigated the profile of IgG and IgG subclass antibody responses to SARS– CoV-2 in Tunisian patients with COVID-19 according to disease severity (86 patients with severe disease and 63 with mild to moderate disease). Two in house developed ELISA with excellent performance were used to test for antibodies to the nucleocapsid (N) protein and the receptor-binding domain of the spike antigen (S-RBD) of SARS-CoV-2. IgG, IgG1 and IgG3 antibodies were significantly higher in patients with severe disease compared to non-severe disease. Antibodies to S-RBD or the N protein were dominated by IgG1 and IgG3 or IgG1/IgG3 and IgG2 subclasses respectively. In patients with severe disease, IgG antibodies ‘ appearance to S-RBD was delayed compared to the N protein. IgG subclass imbalance may reflect the pathophysiology of COVID-19 and may herald disease aggravation. This study brings information on the immune responses to SARS-CoV-2 in North African patients and completes the picture drawn on COVID-19 in different African populations and worldwide.

## 1. Introduction

The emergence and pandemic extension of COVID-19 (Coronavirus disease 2019), caused by SARS-CoV-2 (Severe acute respiratory syndrome coronavirus 2), has led to an unprecedented global public health crisis (https://www.who.int/emergencies/diseases/novel-coronavirus-2019). SARS-CoV-2 is the seventh member of the *Coronaviridae* family of viruses that can infect humans and the third zoonotic coronavirus after SARS-CoV (severe acute respiratory syndrome coronavirus) and MERS-CoV (Middle east respiratory syndrome coronavirus), to cross the species barrier and cause severe respiratory infections in humans. Coronavirus particles contain four main structural proteins, namely the spike (S), membrane (M), envelope (E), and nucleocapsid (N) proteins, all of which are encoded within the 3’ end of the viral genome ***(Fehr and Perlman 2015)***.

The spike (S) and the nucleocapsid (N) proteins are highly immunogenic. The trimeric S protein is the first virus component that binds SARS-CoV-2 to host cell receptors. This interaction occurs through the receptor-binding domain (RBD) of S1 subunit and triggers via the S2 subunit, the fusion of virus to host membranes. The N protein, the most abundant viral protein, is a structural component of the helical nucleocapsid, which plays an important role in viral pathogenesis, replication and RNA packaging ***(McBride et al. 2014)***.

COVID-19 has a broad spectrum of clinical presentations ranging from asymptomatic, pauci-symptomatic, moderate, severe, critical and eventually deadly. Severity of disease may affect the patient ‘s immune response to SARS-CoV-2. Several studies demonstrated that critically ill patients had higher levels of proinflammatory cytokines and lower counts of T lymphocytes than those who have moderate or mild cases ***(Chen et al. 2020a)***. Recent studies found that distinct antibody signatures could characterize different disease outcomes in hospitalized COVID-19 patients. Specifically, Atyeo et al showed that early spike-specific responses were associated with a positive outcome (convalescence), while early N-specific responses were associated with a negative outcome (death) ***(Atyeo et al. 2020)***. In addition, the level of SARS-CoV-2 specific antibodies and neutralizing titers have been shown to be associated with COVID-19 disease severity ***(Long et al. 2020, Garcia-Beltran et al. 2021, Yates et al. 2021, Boonyaratanakornkit et al. 2021, Salazar et al. 2020)***. Few reports investigated IgG subclasses (IgG1, IgG2, IgG3 and IgG4) and their association with disease severity ***(Long et al. 2020, Chakraborty et al. 2021, Dogan et al. 2021, Luo et al. 2021, Yates et al. 2021)***. Thus more work is needed to further characterize the antibody isotypes mainly IgG and subclasses generated in response to SARS-CoV-2 throughout the full clinical spectrum of COVID-19.

Using in-house developed SARS-CoV-2 S-RBD and N-based ELISA assays that demonstrated high sensitivity and specificity, we found that high levels of specific antibodies to S and N were associated with disease severity. In addition we noted a delayed kinetic of anti-S-RBD antibodies that might be predictive of the shift to severe forms of the disease. Our data showed IgG subclass imbalance in association with disease severity with a significantly increased SARS-CoV-2-specific IgG1 and IgG3 levels in severe COVID-19 patients compared to patients with mild to moderate disease.

## 2. Materials and Methods

### 2.1 Study population and serum sampling

A total of 149 individuals with COVID-19 were included in the study: 45 patients admitted at Abderrahmane Mami hospital (Ariana); 63 health workers from Charles Nicolle hospital and Institut Pasteur de Tunis (50 Males; 58 Females; age 25-85 years, median 50 years); To this initial cohort used for assay validation, we added in a second step 41 patients admitted in Yesminette Hospital, Ben Arous (22 Males; 19 Females; age 27-88 years, median 68 years). SARS-CoV-2 was confirmed in all patients by RT-PCR on naso-pharyngeal swabs.

SARS-CoV-2 infected individuals were classified into, *i)* asymptomatic (quarantined and remained asymptomatic), *ii)* mild disease (no sign of pneumonia on imaging, mild clinical symptoms), *iii)* moderate disease (fever, respiratory symptoms and radiological signs of pneumonia without sign of severity (cough, slight dyspnea, FR <30cpm, SpO2> 92%), diarrhea without repercussion), *iv)* severe disease (dyspnea, respiratory rate> 30 / min, blood oxygen saturation <92% in ambient air and / or pulmonary infiltrations> 50%) and *v)* critically ill (respiratory failure, septic shock and / or multiple organ dysfunction or failure or death).

To facilitate data analysis, cases with asymptomatic, mild or moderate disease were classified as “non-severe” (n=63); severely and critically ill patients were classified as “severe” (n= 86).

Blood was collected from patients within the first 6 weeks after onset of illness (n=22 for day 0-7, n=43 for day 8-14, n=30 for day 15-28, n=54 for day 29-42). The serum or plasma were separated and stored at -80°C until use.

Control serum samples (n=72) from healthy donors were collected before the pandemics (between 2013 and 2018).

The study protocol was approved by the Institut Pasteur de Tunis ethical committee (2020/21/I/LR16IPT/V2). Patients were included after written informed consent. Non-inclusion criteria were as follows: presence of a mental handicap, pregnant women and patients on immunosuppressive therapy.

### 2.2 Recombinant proteins production and purification

Recombinant N protein was produced in *Escherichia coli BL21 (DE3)* using the pETM11/N-nCov-(His)6-Nter plasmid as previously described ***(Grzelak et al. 2020b)***. In brief, one selected colony was suspended and cultured in LB medium. When OD_600_ reached 1, iso-propyl-β-D-thiogalactopyranoside (IPTG) was added to a final concentration of 0.2 mM and incubation was continued for 3 h. Subsequently, the bacterial cells were lysed by sonication in the Urea lysis buffer (8M Urea, 100mM NaCl, 20mM Tris-HCl and 10mM Imidazole). The recombinant protein was then purified using affinity chromatography followed by gel filtration. Protein expression and purification was monitored by SDS-polyacrylamide gel electrophoresis.

For proper post-translational modifications (i.e. glycosylation, and correct folding), the S protein and its RBD fragment require to be expressed in eukaryotic systems ***(Farnós et al. 2020, Lv et al. 2020, Perera et al. 2020)***. The S-RBD (residues 319–541) of the SARS-CoV-2 spike protein ***(Wang et al. 2020)*** was cloned into a customized pFastBac vector. Recombinant bacmid DNA was generated using the Bac-to-Bac system following the manufacturer instruction (Life Technologies, Thermo Fisher Scientific).

The recombinant bacmid DNA was transfected into S-f9 cells. Sf-9 cells were then infected with the recombinant baculoviruses at an MOI of 3. 72 h post infection, rRBD protein was collected then purified, after a concentration step, by AKT□ purifier system (GE Healthcare Life Sciences, Uppsala, Sweden) using affinity His Trap Flow fast 5ml column.

The quality control of the produced recombinant proteins was performed using several tests including western blot and MALDI-TOF.

### 2.3 Indirect ELISA for IgG antibodies to S-RBD and N SARS COV 2 proteins

Ninety-six–well ELISA plates were coated overnight with recombinant proteins in phosphate-buffered saline (PBS) (50 ng for N and 100 ng for S-RBD in a volume of 50 µl per well). After washing three times with PBS–0.1% Tween 20 (PBS-T), plates were then incubated with 4% bovine serum albumin in PBST for 1 hour at 37°C for blocking step. After three washes, 50µl of diluted sera (1:200 for the N-based and 1:400 for the S-RBD based assays) in PBS-T was added and incubated for 2 hours at 37°C. Plates were then washed six times, incubated with 8000-fold diluted peroxidase-conjugated goat anti-human IgG (Sigma, AG029) for 1 hour at 37°C. After six washes, plates were revealed by adding 50µl of horseradish peroxidase (HRP) chromogenic substrate [3,3′,5,5′-tetramethylbenzidine (TMB) (BD Biosciences, 555214). After 10 min incubation, the reaction was stopped by adding 50µl of sulfuric acid (2N) and ODs were measured at 450/630 nm.

### 2.4 Indirect ELISA for IgG subclass antibodies to S-RBD and N SARS COV 2 proteins

Plates were coated overnight with recombinant N protein (1µg/ml) or S-RBD (2µg/ml). After blocking with 4% bovine serum albumin in PBS-T for 1 hour at 37°C, the plates were added with serum samples at 1:100 dilution. After washing, plates were added with biotin-conjugated (Sigma) anti-human IgG1 (1:500), anti-human IgG2 (1:7000), anti-human IgG3 (1:6000) and anti-human IgG4 (1:8000) then incubated at 37°C for 1 hour. Subsequently, plates were washed and incubated with streptavidine-horseradish peroxidase (1:15000) for 30 min. After six washes with PBS-T, 50µl of TMB (BD Biosciences, 555214) were added, and plates were incubated for 10 min. The reaction was stopped by adding 50µl of sulfuric acid (2N) and OD was measured at 450/630 nm.

### 2.5 Statistical analysis

Receiving operator characteristic (ROC) curves analysis was performed to determine the accuracy of the in-house ELISA assays considering RT-PCR as the gold standard to define SARS-CoV-2 positivity status. Specificity and sensitivity with their 95% confidence intervals were determined using the optimal cut off values. We further studied agreement between the developed in house ELISA assays and the automated Roche Diagnostics assays by estimating the Kappa coefficient.

Statistical analyses using nonparametric tests were performed using GraphBad Prism version 5 and p<0.05 was considered to be statistically significant.

## 3. Results

### 3.1 Development and validation of N- and S-RBD-based ELISA

The recombinant N protein of SARS-CoV-2 was produced in *E. coli* BL21 (DE3) then purified by affinity chromatography followed by gel filtration to obtain highly purified protein as shown in **Figure 1a**. Quality control assays included MAL-DI-TOF **(data not shown)**. The coding sequence for RBD gene was expressed using SF9 cells then purified. **Figure 1c** and **1d**.

**Figure 1.**
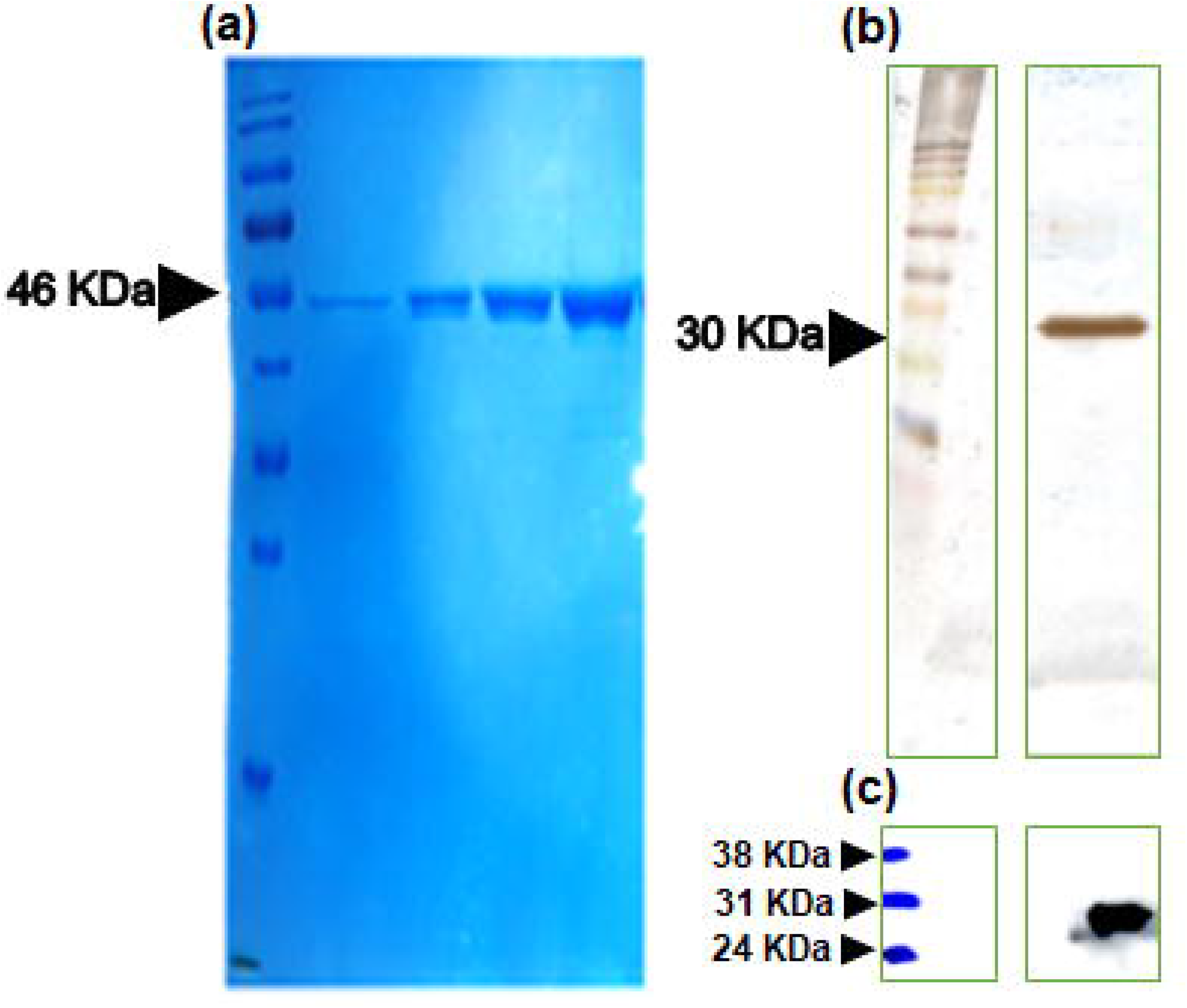
Recombinant proteins expression and purification. Recombinant N and S-RBD proteins were produced in *E*.*coli* BL21 (DE3) and Sf9 cells respectively. **(a)** SDS-PAGE analysis of the N protein purified using Ni-NTA affinity chromatography followed by gel filtration Sephadex G75**; (b)** Silver staining of recombinant S-RBD produced using baculovirus/Sf9 expression system and purified using Ni-NTA affinity followed by gel filtration superdex 200; **(c)** Western blotting using monoclonal anti-His antibody.

ELISA assays for antibodies to the S-RBD and N proteins were subsequently developed and optimized. The performance of both ELISA tests was assessed using a panel of sera from 108 patients with RT PCR confirmed COVID-19 (collected within the first 6 weeks after onset of illness symptoms) and 72 prepandemic sera collected prior to 2018 and cryopreserved within our serobank. As shown in **Figure 2a** and **2d**, COVID-19 sera exhibited significantly higher ODs than prepandemic control sera (p<0.0001). In addition, IgG antibodies reactivity to N and S-RBD proteins were significantly higher in sera collected after day 8 and 15, respectively, compared to those collected earlier **(Fig.2b and 2e)**. The highest levels were obtained with sera collected between day 15 and day 28. The performance of both tests was estimated by the area under the ROC curve (AUC) established using the RT-PCR test as a gold standard. The overall performance of the anti-N and anti-S-RBD ELISA was very high (AUC 0.966 and 0.98, respectively, p< 0.0001) with a sensitivity of 94 % and a specificity of 93% for anti-N test and a sensitivity of 95% and a specificity of 93% for anti-S-RBD test **(Fig.2c, 2f)**. Interestingly, levels of IgG anti-N and IgG anti-S-RBD were positively correlated in all tested patients (Rho= 0.66, p<0.0001, **Fig.3**).

**Figure 2.**
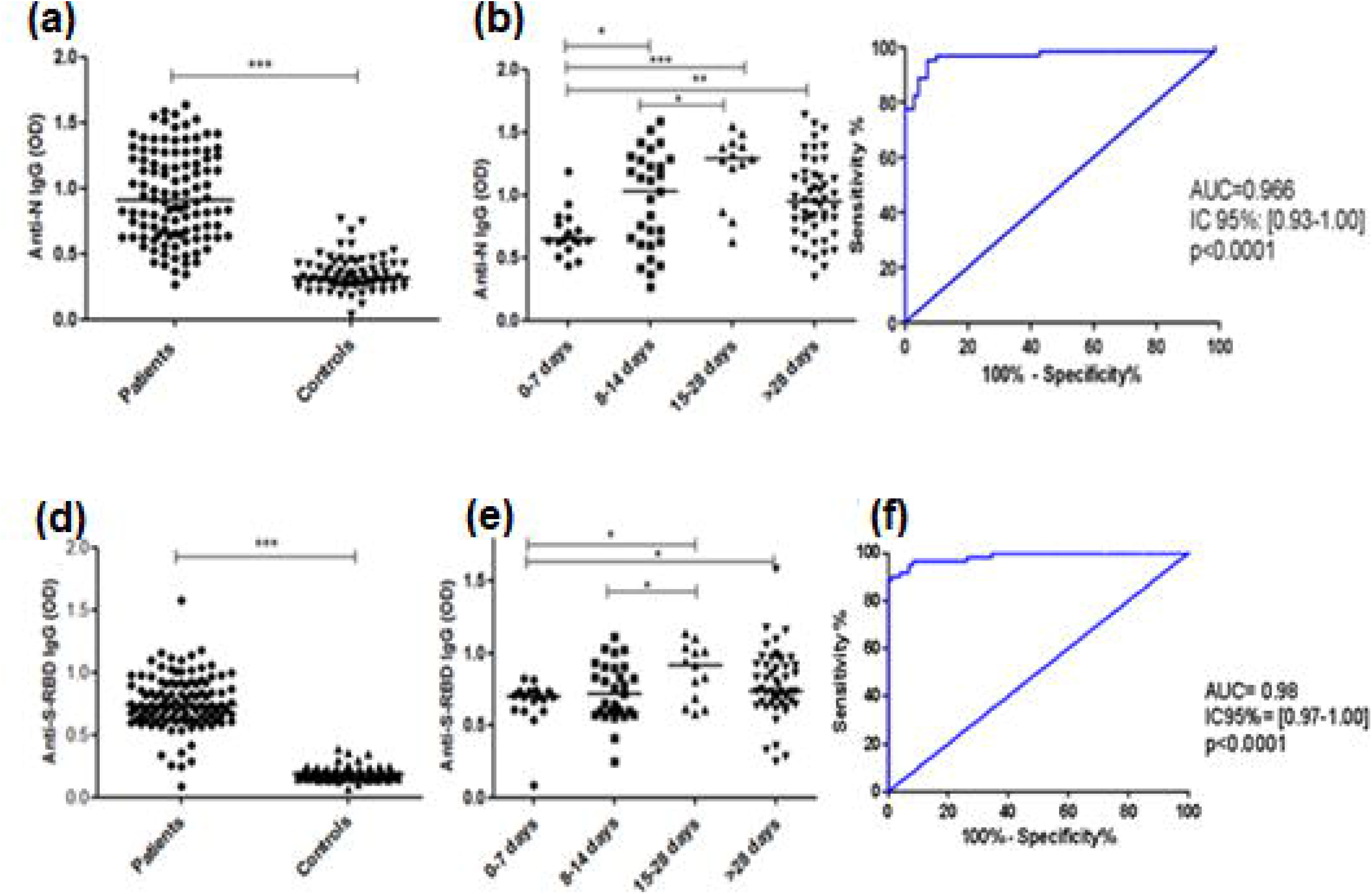
N- and S-RBD-based ELISA assays validation. A total of 108 sera/plasma samples obtained from COVID-19 patients confirmed by PCR and 72 sera samples collected before the pandemics (prior to 2018) from healthy donors were subjected to antibody detection. N-**(a)** and S-RBD **(d)** specific IgG levels are presented in optical densities (OD (450nm-620nm)); N **(b)** and S-RBD **(e)** specific IgG levels obtained in COVID-19 patients were stratified according to the blood collection time after the symptoms onset; ROC curve analysis representing the performance of the N-based **(c)** and S-RBD based **(f)** ELISA tests performances were estimated by the area under the ROC curve (AUC) established using the RT-PCR test as a gold standard. Bars indicate median values. Mann-Whitney U-test was used to compare differences between two groups, a two-tailed p-value <0.05 was considered to be statistically significant. *** p ≤ 0.001, ** p ≤ 0.01 and *p ≤ 0.05.

**Figure 3.**
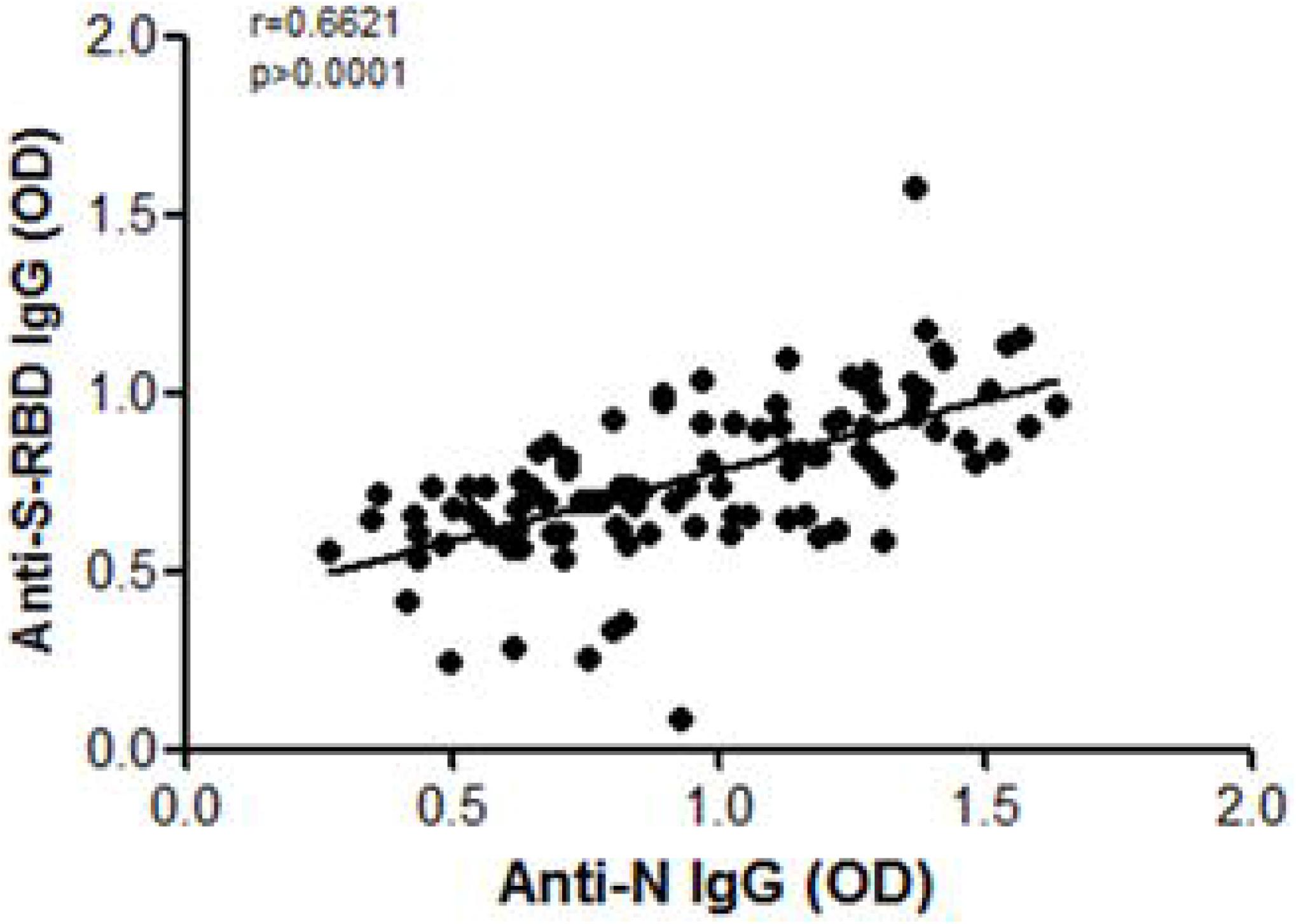
Correlation between IgG antibodies against N and S-RBD proteins in sera of tested COVID-19 patients. Regression line for pairs of optical densities (OD) values corresponding to IgG anti-N and anti S-RBD antibodies is shown for the 108 COVID-19 patients. The Spearman ‘s rank correlation coefficient (rho) and the P value are indicated.

To better evaluate the performances of our ELISA based on N and S-RBD proteins, we did a head-to-head comparison on a set of negative (n=42) and positive cases (n=46) of our tests with the automated Roche Diagnostics. The results of both tests were read blindly to each other. Indeed, it has been demonstrated that both Roche N and Roche S-RBD assays displayed the highest performance automated SARS-CoV-2 serological immunoassays***(Andrey et al. 2021)***. Our assays demonstrated 95%, 93% positive agreement for and 93%, 98% negative percent agreement for the N-based and RBD-based ELISA respectively with Roche Diagnostics, with Cohen ‘s kappa value of 0.88 and 0.9 for N and RBD respectively **(Table 1 and 2)** (strong agreement).

**Table 1.** Head-to-head comparison of the developed anti-N ELISA with the automated Roche N assa

|  |  | Roche N assay |  | Total |
| --- | --- | --- | --- | --- |
|  |  | Negative | Positive |  |
| Anti-N ELISA | Negative | 42 | 2 | 44 |
|  | Positive | 3 | 41 | 44 |
| Total |  | 45 | 43 | 88 |

\*Kappa=0.886

**Table 2:** Head-to-head comparison of the developed anti-S-RBD ELISA with the automated Roche S-RBD assay

|  |  | Roche S-RBD assay |  | Total |
| --- | --- | --- | --- | --- |
|  |  | Negative | Positive |  |
| Anti-S-RBD ELISA | Negative | 42 | 3 | 45 |
|  | Positive | 1 | 42 | 43 |
| Total |  | 43 | 45 | 88 |

\*Kappa=0.909

### 3.2 IgG antibodies to SARS-CoV-2 proteins in association with COVID-19 severity

To better analyze the antibody responses to SARS-CoV-2 proteins across COVID-19 severity groups, we compared two groups with “severe” versus “non severe” clinical presentations. Thus, an additional group of 41 “severe” patients was then added to the cohort of patients. In the total, 86 patients with asymptomatic or mild to moderate cases constituted the group of “non severe” whereas 63 patients with severe and critical cases consisted of the group of “severe” patients.

Our data showed higher levels of both antibodies in “severe” patients compared to “non severe” patients (**Fig.4a)**. The difference was statistically more significant for IgG anti-N antibodies (p<0.0001) than for IgG anti-S-RBD antibodies (p<0.001). The delay for detection of IgG antibodies specific to N and S-RBD after disease symptoms onset was compared between both groups. Interestingly, IgG antibodies to S-RBD appeared later than IgG antibodies to N antigen in the group “severe” disease **(Fig.4b)**. In the “non severe” group, the reactivity of IgG antibodies to the S-RBD remains constant, irrespective of time of symptom onset **(Fig.4c)**.

**Figure 4.**
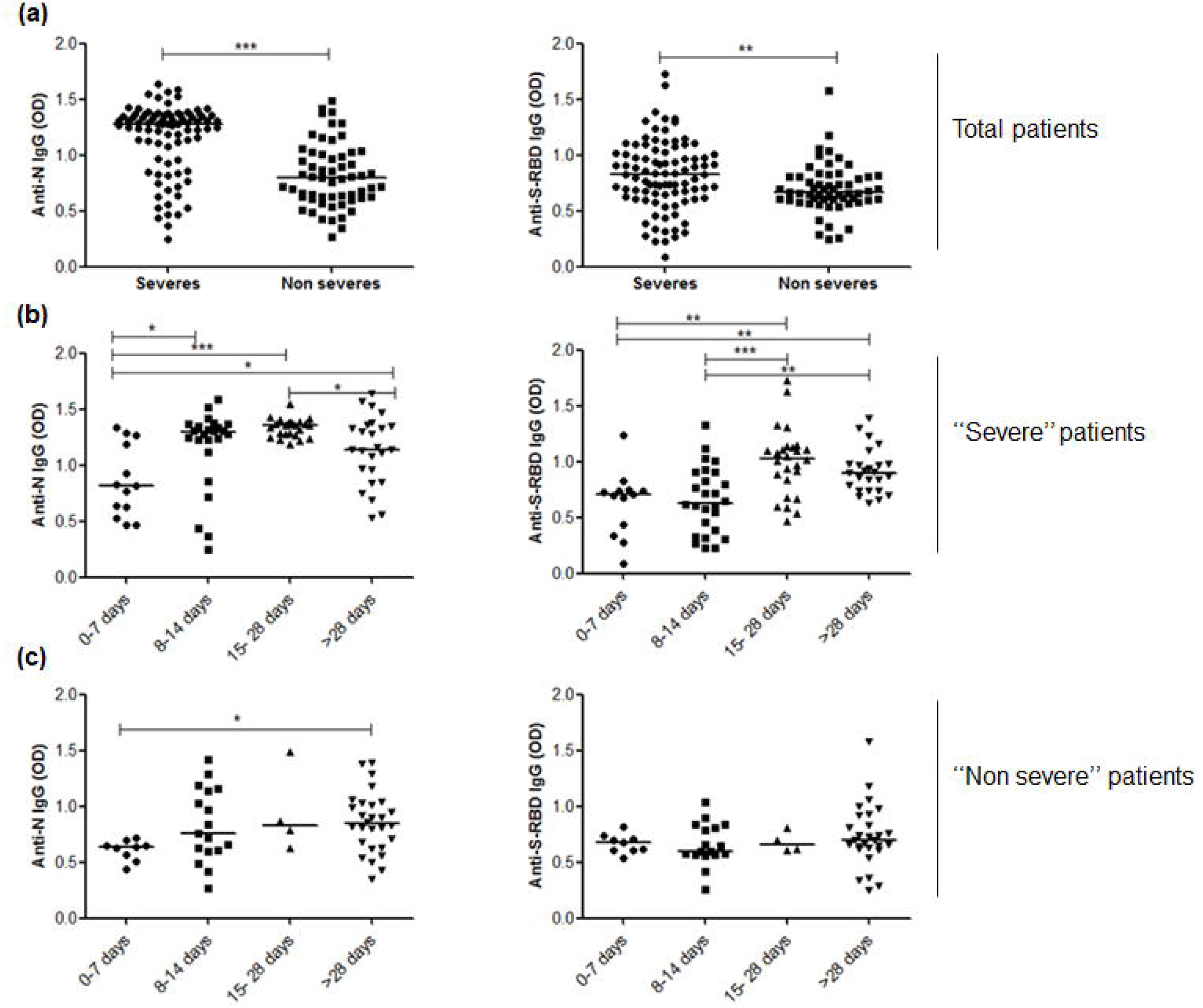
Serum IgG levels of N-and S-RBD specific antibodies in “severe” versus “non severe” COVID-19 patients. IgG anti-N and S-RBD antibodies were assessed in “severe” (n=86) and “non severe” (n=63) patients. **(a)** Optical densities (OD) of IgG anti-N-and S-RBD-proteins obtained in “severe” and “non severe” COVID-19 patients are shown. Bars indicate median values; **(b)** Optical densities (OD) of IgG anti-N-and S-RBD-proteins obtained in “severe” COVID-19 patients stratified according to the collection serum time after disease onset. Bars indicate median values; **(c)** Optical densities (OD) of IgG anti-N-and S-RBD-proteins obtained in “non severe” COVID-19 patients stratified according to the collection serum time after disease onset. Bars indicate median values. Mann-Whitney U-test was used to compare differences between two groups, a two-tailed p-value <0.05 was considered statistically significant. *** p ≤ 0.001, ** p ≤ 0.01 and *p ≤ 0.05.

### 3.3 SARS-CoV-2-specific subclass IgG antibodies in association with COVID-19 disease severity

In humans, IgG immunoglobulins belong to either one of the four functional subclasses, namely IgG1, IgG2, IgG3, and IgG4 that have distinct properties and effectors functions. Thus, we further investigated whether COVID-19 severity was associated with IgG subclass imbalance. Thus, we tested specific IgG antibodies to N and S-RBD for subclass markers in the group of severe disease (n=38) compared to non-severe disease (n=32). Only sera collected over 15 days after onset of illness were analyzed. As shown in **Figure 5a**, specific antibodies to RBD and N antibodies were dominated by IgG1 and IgG3 subclass isotypes. In addition, higher levels IgG1 of IgG3 antibodies were detected in the “severe” group compared to “non severe” ones (p ≤ 0.001). IgG4 antibodies reactive with either N or S-RBD protein were barely detected. However, high levels of IgG2 antibodies to N but not to S-RBD was detected but with no difference between “severe” and “non severe” groups of patients **(Fig.5a)**. Overall, these data indicate some imbalance between IgG subclass antibodies to SARS-CoV-2 proteins, skewed towards a dominant IgG1 and IgG3 response in patients who experienced a severe form of COVID-19. This imbalance is best represented on **Figure 5b** by the ratio of the combined IgG1 and IgG3 antibodies over the combined IgG2 and IgG4 antibodies with a significantly higher ratio in the group of severe disease (p ≤ 0.001).

**Figure 5.**
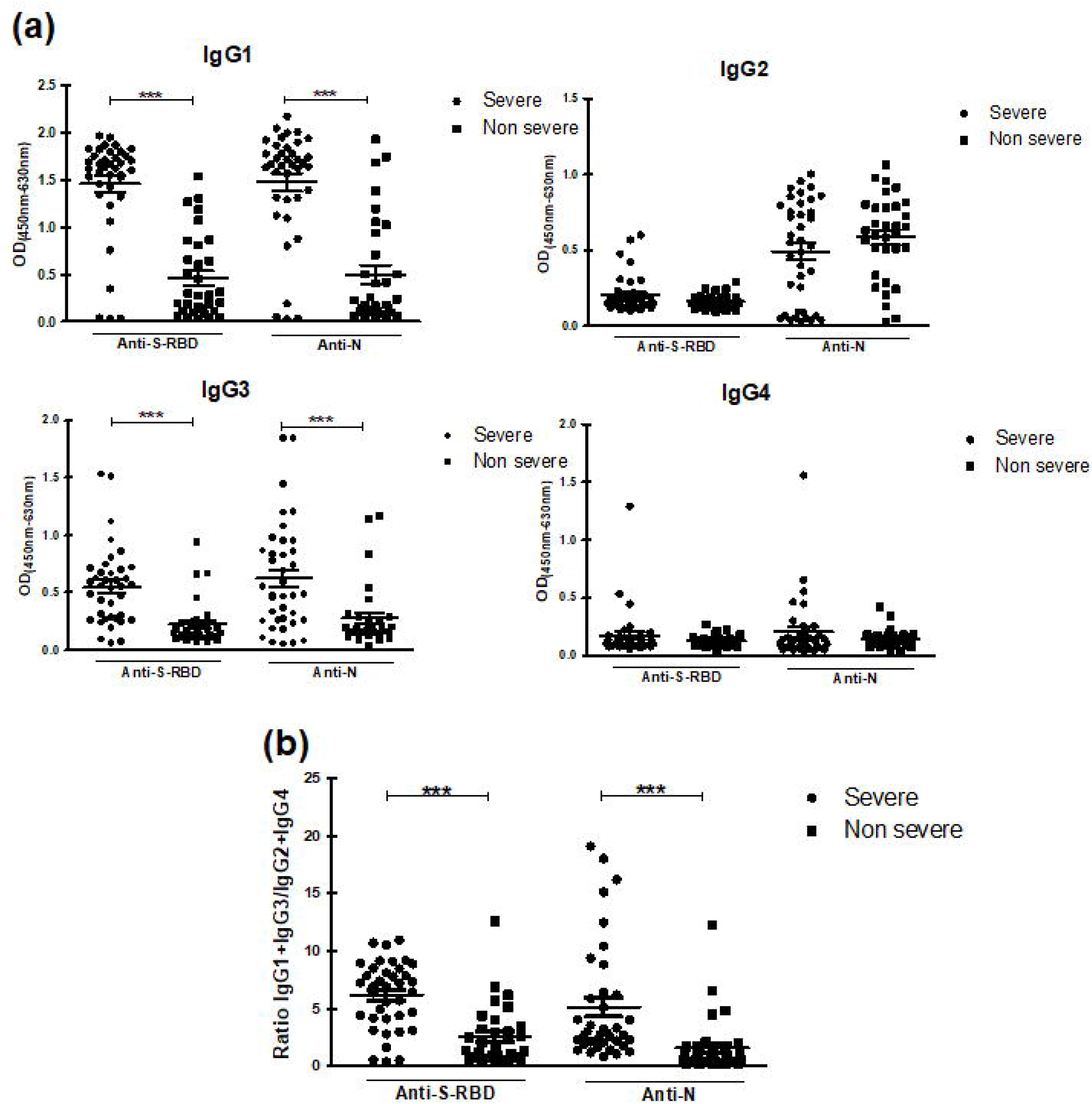
N- and S-RBD-specific IgG subclasses induced in COVID-19 in correlation with disease severity. **(a)** IgG1, IgG2, IgG3 and IgG4 responses to N and S-RBD proteins in “severe” patients (n=38) versus “non severe” patients (n=32) after 15 days from the disease onset; **(b)** Ratio of combined IgG1 and IgG3 response to combined IgG2 and IgG4 response in “severe” and “non severe” patients enrolled after 15 days from the disease onset. OD; optical density. Bars indicate median values. Mann-Whitney U-test was used to compare differences between two groups, a two-tailed p-value <0.05 was considered statistically significant. *** p ≤ 0.001, ** p ≤ 0.01 and *p ≤ 0.05.

## 4. Discussion

During SARS-CoV-2 infection, the nucleocapside (N), the spike (S) and the receptor binding domain of the spike antigen (S-RBD) are immunodominant targets with antibodies to the latter domain playing a major role in neutralizing virus infection of host ‘s cells ***(Premkumar et al. 2020, Smits et al. 2021)***. In addition, these proteins are good targets for serological tests in seroprevalence studies. The latter are essential to assess the true diffusion of SARS-CoV-2 in exposed populations and integrating asymptomatic patients that would otherwise escape detection. To this end, several immuno-enzymatic techniques have been developed, optimized and validated.

ELISA techniques for detection of antibodies to SARS-CoV-2 exhibited similar performance with the S-Flow assay and the luciferase system (LIPS) assay that recognized diverse SARS-CoV-2 antigens in immunoprecipitation ***(Grzelak et al. 2020b)***. In this study, we developed and optimized indirect ELISA assays to detect SARS-CoV-2 specific IgG antibodies in serum samples, using two recombinant viral proteins namely N and S-RBD.

Our in-house-developed ELISA incorporating S-RBD and N recombinant proteins were able to efficiently discriminate COVID-19 patients from healthy control as assessed by the ROC curve analysis. The overall performance of the anti-N and anti-S-RBD ELISA assays was very high (AUC: 0.966 and 0.98, respectively, p< 0.0001) with a sensitivity of 94%/ 93% and a specificity of 95%/93% for the anti-N and -S-RBD tests, respectively. The developed ELISA displayed a strong agreement with both Roche N and S-RBD, a commercial kit that is recognized as giving the highest performances among automated serological immunoassays of SARS-CoV-2 (Andrey et al. 2021). In addition, using the developed tests, we showed a relatively good correlation between N and S-RBS antibodies in COVID-19 patients in the first 6 weeks post infection with r =0.66, a result corroborated by data from Grzelak et al ***(Chen et al. 2020b, Grzelak et al. 2020a)***.

In the present study, we used the in house developed assays to investigate whether disease severity was associated with SARS-CoV-2 specific immunoglobulin G (IgG) and subclass isotypes. To this end, sera from 86 patients with “severe” COVID-19 and 63 patients with “non severe” COVID-19 allowed us to analyze the SARS-CoV-2 antibody response in patients with different levels of disease severity. We found that, 2-4 weeks after onset symptoms, patients with severe disease mount a robust humoral immune response against either N or S-RBD antigens of SARS-CoV-2. In contrast, mild infections show significantly lower antibody levels. Such data are in line with previous studies conducted in populations with diverse ethnicity reporting more robust humoral immunity in recovered patients who had symptomatic COVID-19 compared to asymptomatic ones ***(Grzelak et al. 2020a, Lucas et al. 2020, Andrey et al. 2021, Dogan et al. 2021, Mehdi et al. 2021, Yates et al. 2021)***. The lower level of IgG responses associated with mild disease may reflect lower viral loads and viral antigens. Interestingly, a recent study suggests that most severely affected patients had not only the highest anti-S-RBD and anti-spike antibody levels but also the highest levels of inflammatory markers and pro-inflammatory cytokine signatures ***(Chakraborty et al. 2021)***. Moreover, patients who received corticosteroids had decreased levels of antibodies to S-RBD IgG as well as lower neutralization titers ***(Garcia-Beltran et al. 2021)***. Our data also indicate a delay in the kinetics of appearance of IgG anti-S-RBD antibodies in “severe” patients when compared to that of IgG antibodies to the N protein. In the “non severe” group, the level of antibodies to the S-RBD protein remains constant independently of time of symptom onset. Consistently, Lucas et al. showed that patients who ultimately died of COVID-19 mount a robust, yet delayed, response of anti-S, anti-S-RBD IgG and neutralizing antibody (NAb) levels compared to survivors ***(Lucas et al. 2020)***. Delayed seroconversion kinetics correlated with impaired viral control in deceased patients ***(Lucas et al. 2020)***. Thus, a delay in the appearance of neutralizing antibodies in some subjects associated with the early appearance of high titers of anti-N antibodies (which might be facilitating antibodies) could be predictive of the shift to severe forms of the disease. This suggests that non-neutralizing antibodies likely play a pathogenic role. Indeed, in contrast to the protective role of neutralizing antibodies, the immune system may produce antibodies that are not able to inhibit the replication cycle, nor mediate virus clearance and elimination of virus-infected cells through antibody-dependent cellular cytotoxicity and phagocytosis. Actually, such antibodies would mediate enhanced virus cell entry and induction of severe inflammatory activation through antibody-dependent enhancement (ADE) ***(Taylor et al. 2015, Arvin et al. 2020, Kulkarni 2020)***.

In humans, the four IgG subclasses IgG1, IgG2, IgG3 and IgG4 exhibit different functional activities. For instance, IgG1 and IgG3 antibodies are generally induced in response to protein antigens and typify Th1 responses while IgG2 and IgG4 antibodies are mainly associated with polysaccharide antigens and typify Th2 responses ***(Schroeder and Cavacini 2010)***.

In viral infections, IgG1 and IgG3 antibodies are the predominant IgG subclasses and are well reported to provide antiviral activity. Both of them are involved in neutralization, opsonization, complement fixation and antibody-dependent cellular cytotoxicity (ADCC) ***(Wu et al. 2020, Garcia-Beltran et al. 2021)***. Such subclasses were recently associated with SARS-CoV-2 infection ***(Suthar et al. 2020, Dogan et al. 2021, Mazzini et al. 2021, Yates et al. 2021)***. Our results are in line with these previous findings and confirm the increase of IgG1 and IgG3 subclasses in patients with “severe” disease compared to patients with “non severe”disease. IgG1 and IgG3 predominance was demonstrated for either antibodies to N or S-RBD. More importantly, Yates et. al demonstrated that spike-specific IgG1 antibodies but not IgG3 were closely correlated with *in vitro* viral neutralization ***(Yates et al. 2021)***.

In contrast to other reports ***(Luo et al. 2021, Mazzini et al. 2021)***, our data show that the antibody responses to the N protein are also dominated by IgG2 subclasses and could thus reflect a mixed Th1/Th2 response. Surprisingly, no significant difference was found between IgG2 anti-N antibody response between “severe” and “non severe” patients. Finally, IgG4 was barely detected in COVID-19 patients as reported by others ***(Luo et al. 2021, Yates et al. 2021)***.

Our study has some limitations. We did not evaluate the effects of gender and age on antibodies responses in association with COVID-19 severity nor did we test to what extent comorbidities may impact the immune response against SARS-CoV-2. Further longitudinal studies are needed to monitor how the immune responses of patients with COVID-19 evolve over time in order to assess the immune signatures that may herald either recovery or aggravation.

## 5. Conclusion

Our study brings information on the immune responses to SARS-CoV-2 in Tunisian patients and points out the fact that IgG subclass imbalance may not only reflect the pathophysiology of COVID-19, but also herald disease aggravation.

The excellent performances of the in house-developed ELISA assays made possible their wider use in seroepidemiological studies across Africa, being conducted within a collaborative project on COVID-19 pandemic led by a consortium of ten African countries. This effort would complete the picture drawn on COVID-19 in different African populations.

## Data Availability

All data produced in the present study are available upon reasonable request to the authors

## Author Contributions

Conception and design of the study was accounted by C.B. and M.B.A. Performed experiments by S.M., W.B.H., K.T., M.B., S.B.H., R.O., S.B. and A.C. Samples were provided by Y.G., I.S., S.Y., J.B.K., A.H., M.A., Y.C., S.P. N.E. and C.K.P.M. Analyzed and interpreted the data with valuable intellectual input by C.B., M.B.A., J.B., S.R., M.R.B. and K.D. Drafting the manuscript was done by C.B. and M.B.A. Manuscript was revised by C.B., S.M., K.T., M.B., W.B.H., S.Q., K.D., J.B., M.R.B. and M.B.A. All authors approved the submitted version.

### Ethics approval Statement

The study protocol was approved by the Institut Pasteur de Tunis ethical committee (2020/21/I/LR16IPT/V2).

### Patient Consent Statement

Informed consent was obtained from all subjects involved in the study.

### Data Availability Statement

Not applicable.

### Permission to reproduce material from other sources

Not applicable.

#### Abbreviations

ADCC: Antibody-dependent cellular cytotoxicity
ADE: Antibody-dependent enhancement
AUC: Area under curve
COVID-19: Coronavirus disease 2019
*E. coli:*: *Escherichia coli*
ELISA: Enzyme-Linked Immunosorbent Assay
HRP: Horseradish peroxidase
LB: Luria Broth
LIPS: Luciferase immunoprecipitation system
MALDI-TOF: Matrix assisted laser desorption ionization-time of flight
N: Nucleocapsid protein
PBS: Phosphate buffered saline
ROC: Receiver operating curve
RT-PCR: Reverse transcription polymerase chain reaction
SARS-CoV-2: Severe acute respiratory syndrome coronavirus 2
S-RBD: Receptor-binding domain of the spike protein

## Acknowledgments

We thank people who consented to participate in this study. We also thank Dr. Sinda Zarrouk for her help finding some material donation.

